# Deciphering causal proteins in Alzheimer’s disease: A novel Mendelian randomization method integrated with AlphaFold3 for 3D structure prediction

**DOI:** 10.1101/2023.02.20.23286200

**Authors:** Minhao Yao, Gary W. Miller, Badri N. Vardarajan, Andrea A. Baccarelli, Zijian Guo, Zhonghua Liu

## Abstract

Hidden confounding biases hinder identifying causal protein biomarkers for Alzheimer’s disease in non-randomized studies. While Mendelian randomization (MR) can mitigate these biases using protein quantitative trait loci (pQTLs) as instrumental variables, some pQTLs violate core assumptions, leading to biased conclusions. To address this, we propose MR-SPI, a novel MR method that selects valid pQTL instruments using the Anna Karenina Principle and performs robust post-selection inference. Integrating MR-SPI with AlphaFold3, we developed a computational pipeline to identify causal protein biomarkers and predict 3D structural changes. Applied to genome-wide proteomics data from 54,306 UK Biobank participants and 455,258 subjects (71,880 cases and 383,378 controls) for a genome-wide association study of Alzheimer’s disease, we identified seven proteins (TREM2, PILRB, PILRA, EPHA1, CD33, RET, and CD55) with structural alterations due to missense mutations. These findings offer insights into the etiology and potential drug targets for Alzheimer’s disease.

## 1. Introduction

Alzheimer’s disease (AD) stands as the primary cause of dementia globally, exerting a considerable strain on healthcare resources^1,2^. Despite extensive efforts, the etiology and pathogenesis of AD are still unclear, and strategies aimed at impeding or delaying its clinical advancement have largely remained challenging to achieve^1,3,4^. The amyloid cascade hypothesis posits that AD begins with the accumulation and aggregation of amyloid-beta (Aβ) peptides in the brain, culminating in the formation of β-amyloid fibrils, leading to tau hyperphosphorylation, neurofibrillary tangle formation and neurodegeneration^5,6^. However, current AD therapies targeting Aβ production and amyloid formation offer only transient symptomatic relief and fail to halt disease progression, resulting in a lack of effective drugs for AD^1,7^. Therefore, it is imperative and urgent to identify causal protein biomarkers to elucidate the underlying mechanisms of AD, and to expedite the development of effective therapeutic interventions for AD.

In causal inference, randomized controlled trials (RCTs) serve as the gold standard for evaluating the causal effect of an exposure on the health outcome of interest. However, it might be neither feasible nor ethical to perform RCTs where protein levels are considered as the exposures. Mendelian randomization (MR) leverages the random assortment of genes from parents to offspring to mimic RCTs to establish causality in non-randomized studies^8-10^. MR uses genetic variants, typically single-nucleotide polymorphisms (SNPs), as instrumental variables (IVs) to assess the causal association between an exposure and a health outcome^11^. Recently, many MR methods have been developed to investigate causal relationships using genome-wide association study (GWAS) summary statistics data that consist of effect estimates of SNP-exposure and SNP-outcome associations from two sets of samples, which are commonly referred to as the two-sample MR designs^12-15^. Since summary statistics are often publicly available and provide abundant information of associations between genetic variants and complex traits/diseases, two-sample MR methods become increasingly popular^14,16-18^. In particular, recent studies with large-scale proteomics data have unveiled numerous protein quantitative traits loci (pQTLs) associated with thousands of proteins^19,20^, facilitating the application of two-sample MR methods, where pQTLs serve as IVs and protein levels serve as exposures, to identify proteins as causal biomarkers for complex traits and diseases.

To employ MR for identifying causal protein biomarkers, conventional MR methods require the pQTLs included in the analysis to be valid IVs for reliable causal inference. A pQTL is called a valid IV if the following three core IV assumptions hold^9,21^:

(A1). **Relevance**: The pQTL is associated with the protein;
(A2). **Effective Random Assignment**: The pQTL is not associated with any unmeasured confounder of the protein-outcome relationship; and
(A3). **Exclusion Restriction**: The pQTL affects the outcome only through the protein in view.

Among the three core IV assumptions (A1) - (A3), only the first assumption (A1) can be tested empirically by selecting pQTLs significantly associated with the protein. However, assumptions (A2) and (A3) cannot be empirically verified in general and may be violated in practice, which may lead to a biased estimate of the causal effect. For example, violation of (A2) may occur due to the presence of population stratification^9,22^; and violation of (A3) may occur in the presence of horizontal pleiotropy^9,23^, which is a widespread biological phenomenon that the pQTL IV affects the outcome through other biological pathways that do not involve the protein in view, for example, through alternative splicing or micro-RNA effects^24-26^.

Recently, several MR methods have been proposed to handle invalid IVs under certain assumptions, as summarized in Table 1. Some of these additional assumptions for the identification of the causal effect in the presence of invalid IVs are listed below:

**Main Table 1:**
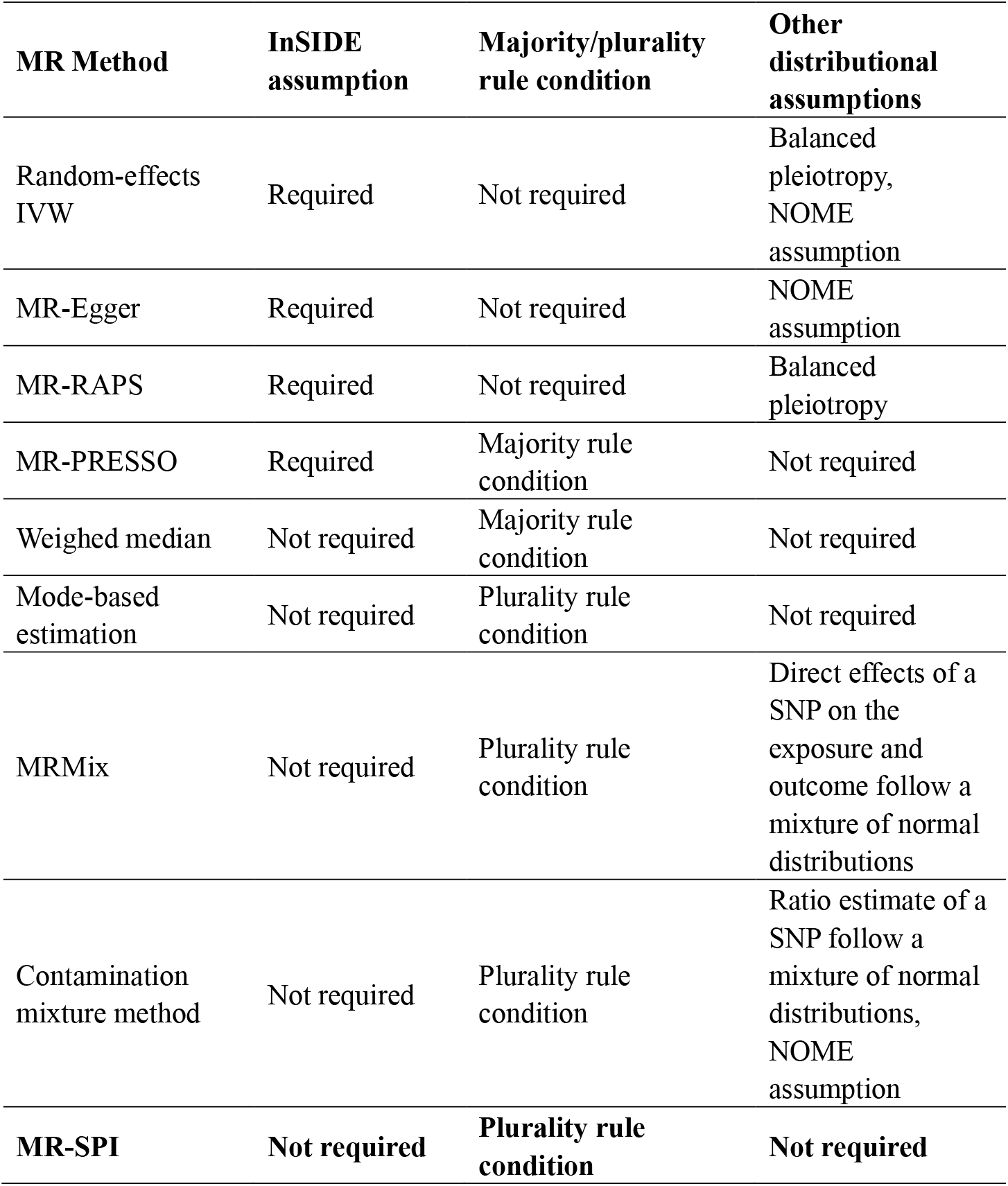
Comparison of MR methods and the underlying assumptions for handling invalid IVs. Balanced pleiotropy means on average the pleiotropic effects have zero mean. NOME assumption refers to NO Measurement Error in the exposure data.

i. The Instrument Strength Independent of Direct Effect (InSIDE) assumption: the pQTL-protein effect is asymptotically independent of the horizontal pleiotropic effect when the number of pQTL IVs goes to infinity. For example, the random-effects inverse-variance weighted (IVW) method^27^, MR-Egger^28^, MR-RAPS (Robust Adjusted Profile Score)^16^, and the Mendelian randomization pleiotropy residual sum and outlier (MR-PRESSO) test^29^.
ii. Majority rule condition: up to 50% of the candidate pQTL IVs are invalid. For example, the weighted median method^30^ and MR-PRESSO.
iii. Plurality rule condition or the ZEro Modal Pleiotropy Assumption (ZEMPA)^15,31^: a plurality of the candidate IVs are valid, which is weaker than the majority rule condition. For example, the mode-based estimation^31^, MRMix^32^ and the contamination mixture method^33^.
iv. Other distributional assumptions. For example, MRMix and the contamination mixture method impose normal mixture distribution assumption on the genetic associations and the ratio estimates, respectively.

Despite many existing efforts, current MR methods still face new challenges when dealing with pQTLs IVs for analyzing proteomics data. First, it’s worth noting that the number of pQTLs for each protein tends to be small. For example, in two proteomics studies, the median number of pQTLs per protein is 4^20,34^. With such a limited number of IVs, those MR methods based on the InSIDE assumption which requires a large number of IVs or other distributional assumptions might yield unreliable results in the presence of invalid IVs^15,28^. Second, current MR methods require an ad-hoc set of pre-determined genetic IVs, which is often obtained by selecting genetic variants with strong pQTL-protein associations in proteomics data^35^. Since such traditional way of selecting IVs only requires the proteomics data, hence the same set of selected IVs is used for assessing the causal relationships between the protein in view and different health outcomes. Obviously, this one-size-fits-all strategy for selecting IVs might not work well for different outcomes because the underlying genetic architecture may vary across outcomes. For example, the pattern of horizontal pleiotropy might vary across different outcomes. Therefore, it is desirable to develop an automatic and computationally efficient algorithm to select a set of valid genetic IVs for a specific protein-outcome pair to perform reliable causal inference, especially when the number of candidate pQTL IVs is small.

In this paper, we develop a novel all-in-one pipeline for causal protein biomarker identification and 3D structural alteration prediction using large-scale genetics, proteomics and phenotype/disease data, as illustrated in Figure 1. Specifically, we propose a two-sample MR method and algorithm that can automatically <u>S</u>elect valid pQTL IVs and then performs robust <u>P</u>ost-selection <u>I</u>nference (MR-SPI) for the causal effect of proteins on the health outcome of interest. The key idea of MR-SPI is based on the Anna Karenina Principle which states that all valid instruments are alike, while each invalid instrument is invalid in its own way – paralleling Leo Tolstoy’s dictum that “all happy families are alike; each unhappy family is unhappy in its own way”^36^. In other words, valid instruments will form a group and should provide similar ratio estimates of the causal effect, while the ratio estimates of invalid instruments are more likely to be different from each other. With the application of MR-SPI, we can not only identify the causal protein biomarkers associated with disease outcomes, but can also obtain missense genetic variations (used as pQTL IVs) for those identified causal proteins. These missense pQTL IVs will induce changes of amino acids, leading to 3D structural changes of these proteins. A classic example of a missense mutation was found in sickle cell disease, where the mutation at SNP rs334, located on chromosome 11 (11p15.4), results in the change of codon 6 of the beta globin chain from [GAA] to [GTA] ^37-39^. This substitution leads to the replacement of glutamic acid with valine at position 6 of the beta chain of the hemoglobin protein, altering the structure and function of hemoglobin protein. Consequently, red blood cells assume a crescent or sickle-shaped morphology, impairing blood flow to various parts of the body^40,41^. Moreover, to further offer novel biological insights into the interpretation of the causal effect at the molecular level, we incorporate AlphaFold3^42-45^ into our pipeline to predict the 3D structural alteration resulting from the corresponding missense pQTL IVs for the causal proteins identified by MR-SPI. Our pipeline can elucidate the mechanistic underpinnings of how missense genetic variations translate into 3D structural alterations at the protein level, thereby advancing our understanding of disease etiology and potentially informing targeted therapeutic interventions.

**Main Figure 1.**
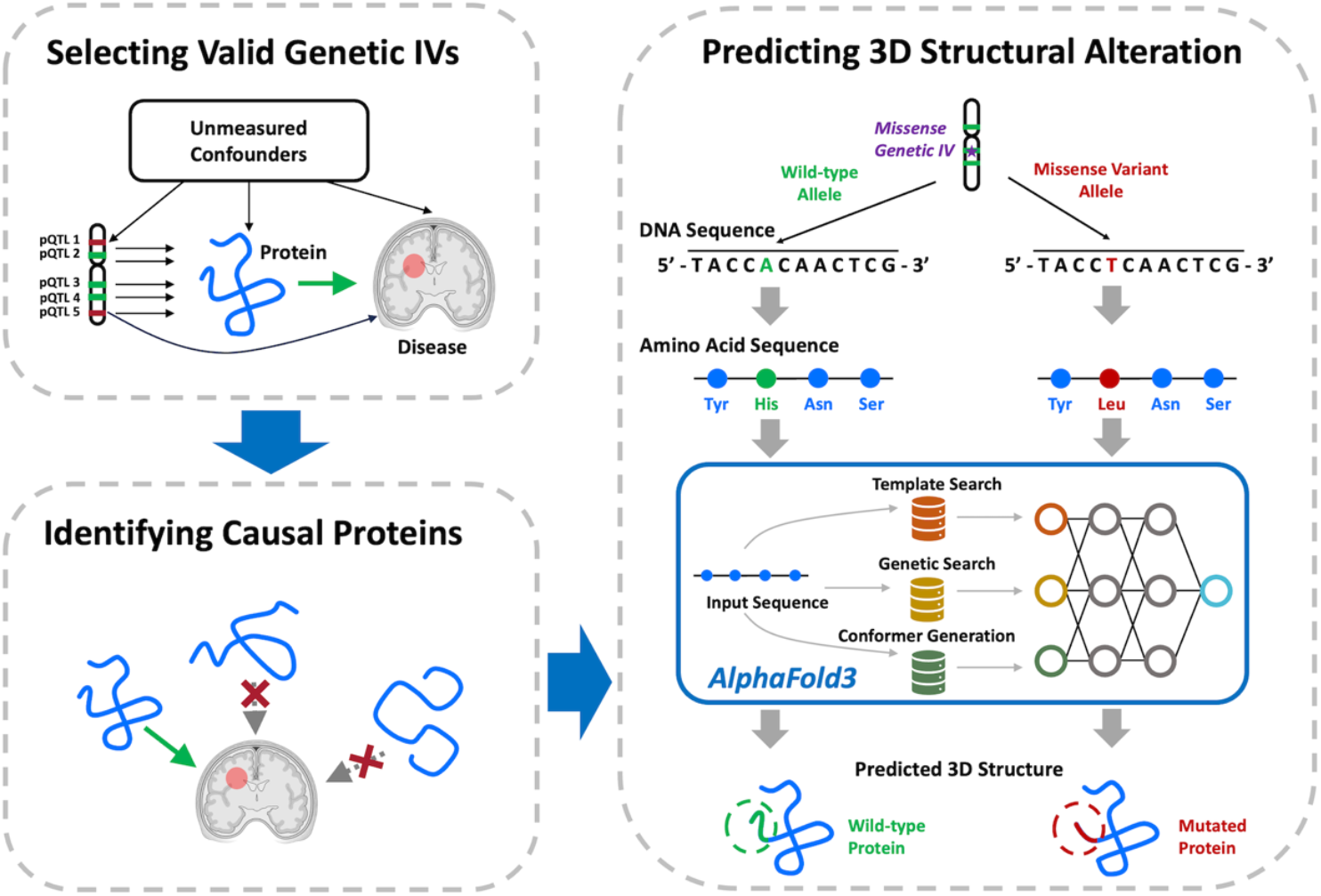
Overview of the pipeline. First, we apply MR-SPI for each protein to (1) select valid pQTL IVs under the plurality condition, and (2) estimate the causal effect on the outcome of interest. Second, we perform the Bonferroni correction procedure for causal protein identification. Third, for each causal protein biomarker, we apply AlphaFold3 to predict the 3D structural alterations due to missense pQTL IVs selected by MR-SPI.

Our pioneering pipeline for the first time integrates the identification of causal protein biomarkers for health outcomes and the subsequent analysis of their 3D structural alterations into a unified framework, leveraging increasingly publicly available GWAS summary statistics for health research. Within our framework, the proposed MR-SPI serves dual purposes: (1) identifying causal protein biomarkers; and (2) selecting valid missense pQTL IVs for subsequent 3D structural analysis. Compared to existing two-sample MR methods, MR-SPI is the first MR method that utilizes both exposure and outcome data to automatically select a set of valid IVs, especially when the number of candidate IVs is small in proteomics data, which is a prominent challenge with no satisfactory solution up to date. We note that while MR-PRESSO also selects valid IVs for MR analysis, it requires both the stronger majority rule condition and the InSIDE assumption, as well as a minimum number of four candidate IVs for implementation^29^. In contrast, our proposed MR-SPI does not require the InSIDE assumption and only requires the plurality rule condition that is weaker than the majority rule condition, and only requires a minimum number of three IVs for the proposed voting procedure. Therefore, MR-SPI is more suitable for analyzing proteomics data. Extensive simulations show that our MR-SPI method outperforms other competing MR methods under the plurality rule condition. We employ MR-SPI to perform omics MR (xMR) with 912 plasma proteins using the large-scale UK Biobank proteomics data in 54,306 UK Biobank participants^20^ and find 7 proteins significantly associated with the risk of Alzheimer’s disease. We further use AlphaFold3^42-45^ to predict the 3D structural alterations of these 7 proteins due to missense genetic variations, and then illustrate the structural alterations graphically using the PyMOL software (https://pymol.org), providing new biological insights into their functional roles in AD development and may aid in identifying potential drug targets.

## 2. Results

### 2.1 Overview of the pipeline

Our proposed all-in-one pipeline for the identification and 3D structural alteration prediction of causal protein biomarkers consists of three primary steps, as illustrated in Figure 1. First, for each protein biomarker, we employ our proposed MR-SPI to select valid pQTL IVs by incorporating the proteomics GWAS and disease outcome GWAS summary data together, and then estimate the causal effect of each protein on the outcome using the selected valid pQTL IVs. The main idea and more detailed implementation steps for MR-SPI is described in Section 2.2. Second, we perform Bonferroni correction^46^ for the *p*-values of the estimated causal effects to identify putative causal protein biomarkers associated with the outcome. Third, for each identified protein, we apply AlphaFold3 to predict and compare the 3D structures of both the wild-type protein and mutated protein resulting from missense pQTL IVs.

### 2.2 MR-SPI selects valid genetic instruments by a voting procedure

MR-SPI is an automatic procedure to select valid pQTL instruments and perform robust causal inference using two-sample GWAS and proteomics data. In summary, MR-SPI consists of the following four steps, as illustrated in Figure 2:

**Main Figure 2.**
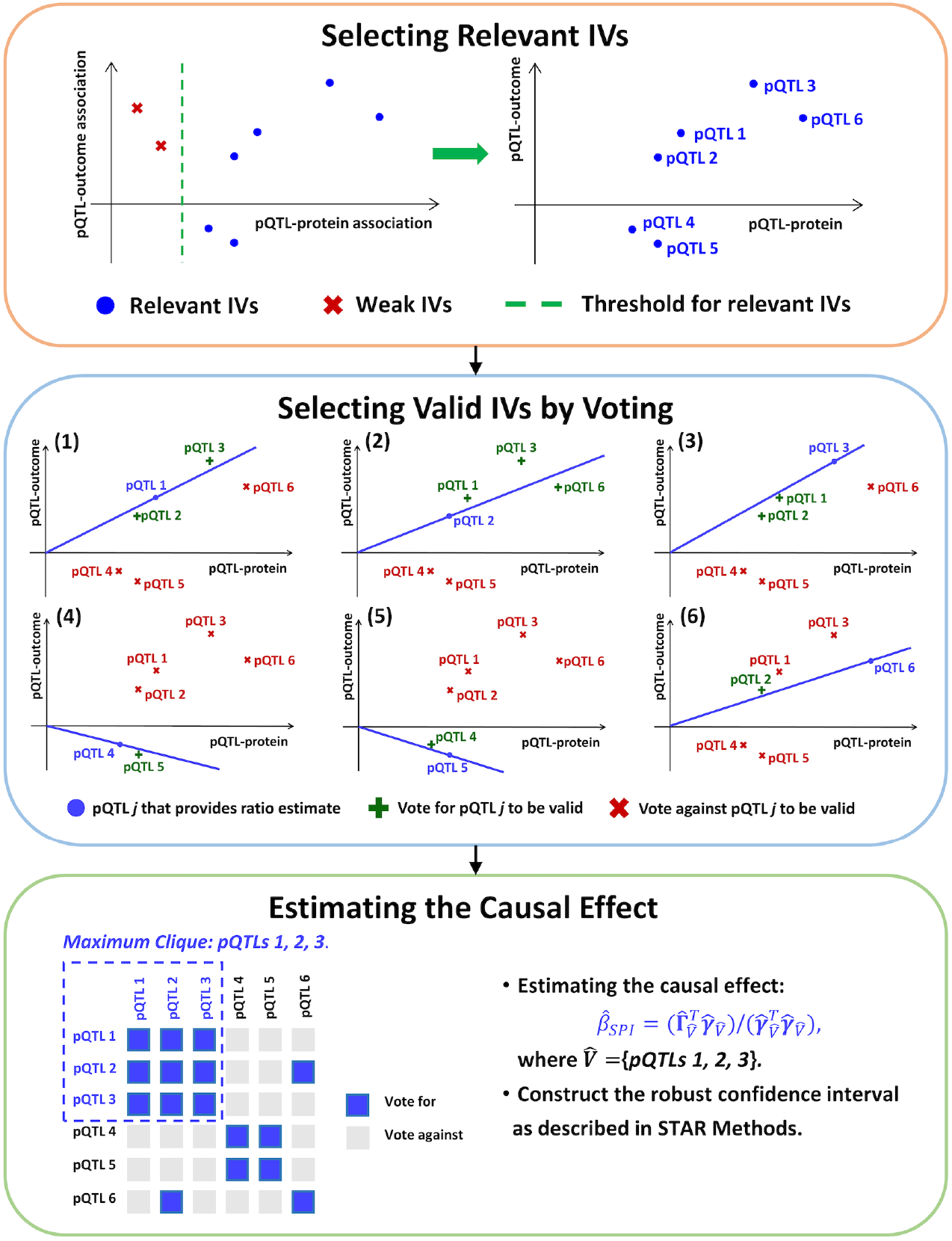
The MR-SPI framework. First, MR-SPI selects relevant IVs with strong pQTL-protein associations. Second, each relevant IV provides a ratio estimate of the causal effect and then receives votes on itself to be valid from the other relevant IVs whose degrees of violation of (A2) and (A3) are small under this ratio estimate of causal effect. For example, by assuming pQTL 1 is valid, the slope of the line connecting pQTL 1 and the origin represents the ratio estimate of pQTL 1, and pQTLs 2 and 3 vote for pQTL 1 to be valid because they are close to that line, while pQTLs 4, 5 and 6 vote against it since they are far away from that line. Third, MR-SPI estimates the causal effect by fitting a zero-intercept OLS regression of pQTL-outcome associations on pQTL-protein associations and construct the robust confidence interval using selected valid pQTL IVs in the maximum clique of the voting matrix, which encodes whether two pQTLs mutually vote for each other to be valid IVs.

1. select relevant pQTL IVs that are strongly associated with the protein;
2. each relevant pQTL IV provides a ratio estimate of the causal effect, and then all the other relevant pQTL IVs votes for it to be a valid IV if their degrees of violation of assumptions (A2) and (A3) are smaller than a data-dependent threshold as in equation (4);
3. select valid pQTL IVs by majority/plurality voting or by finding the maximum clique of the voting matrix that encodes whether two relevant pQTL IVs mutually vote for each other to be valid (the voting matrix is defined in equation (6) in STAR (<u>s</u>tructured, transparent, <u>a</u>ccessible <u>r</u>eporting) Methods);
4. estimate the causal effect using the selected valid pQTL IVs and construct a robust confidence interval with guaranteed nominal coverage even if in the presence of possible IV selection error in finite samples.

Most current two-sample MR methods only use step (1) to select (relevant) pQTL instruments for downstream MR analysis, while the selected pQTL instruments might violate assumptions (A2) and (A3), leading to possibly unreliable scientific findings. To address this issue, MR-SPI automatically select valid pQTL instruments for a specific protein-outcome pair by further incorporating the outcome GWAS data. Our key idea of selecting valid pQTL instruments is that, under the plurality rule condition, valid IVs will form the largest group and should give “similar” ratio estimates according to the Anna Karenina Principle (see STAR Methods). More specifically, we propose the following two criteria to measure the similarity between the ratio estimates of two pQTLs *j* and *k* in step (2):

C1 We say the *k* th pQTL “votes for” the *j* th pQTL to be a valid IV if, by assuming the *j* th pQTL is valid, the *k* th pQTL’s degree of violation of assumptions (A2) and (A3) is smaller than a data-dependent threshold as in equation (4);
C2 We say the ratio estimates of two pQTLs *j* and *k* are “similar” if they mutually vote for each other to be valid.

In step (3), we construct a symmetric binary voting matrix to encode the votes that each relevant pQTL receives from other relevant pQTLs: the (*k, j*) entry of the voting matrix is 1 if pQTLs *j* and *k* mutually vote for each other to be valid, and 0 otherwise. We propose two ways to select valid pQTL IVs based on the voting matrix (see STAR Methods): (1) select relevant pQTLs who receive majority voting or plurality voting as valid IVs; and (2) use pQTLs in the maximum clique of the voting matrix as valid IVs^47^. Our simulation studies show that the maximum clique method can empirically offer lower false discovery rate (FDR)^48^ and higher true positive proportion (TPP) as shown in Table S4 and Supplementary Section S6.

In step (4), we estimate the causal effect by fitting a zero-intercept ordinary least squares regression of pQTL-outcome associations on pQTL-protein associations using the set of selected valid pQTL IVs, and then construct a standard confidence interval for the causal effect using standard linear regression theory. In finite samples, some invalid IVs with small (but still nonzero) degrees of violation of assumptions (A2) and (A3) might be incorrectly selected as valid IVs, commonly referred to as “locally invalid IVs”^49^. To address this possible issue, we propose to construct a robust confidence interval with a guaranteed nominal coverage even in the presence of IV selection error in finite-sample settings using a searching and sampling method^49^, as described in Supplementary Figure S17 and STAR Methods.

### 2.3 Comparing MR-SPI to other competing MR methods in simulation studies

We conduct extensive simulations to evaluate the performance of MR-SPI in the presence of invalid IVs. We simulate data in a two-sample setting under four setups: (**S1**) majority rule condition holds, and no locally invalid IVs exist; (**S2**) plurality rule condition holds, and no locally invalid IVs exist; (**S3**) majority rule condition holds, and locally invalid IVs exist; (**S4**) plurality rule condition holds, and locally invalid IVs exist. More detailed simulation settings are described in STAR Methods. We compare MR-SPI to the following competing MR methods: (1) the random-effects IVW method^27^, (2) MR-RAPS^16^, (3) MR-PRESSO^29^, (4) the weighted median method^30^, (5) the mode-based estimation^31^, (6) MRMix^32^, and (7) the contamination mixture method^33^. We exclude MR-Egger in this simulation since it is heavily biased in our simulation settings. For simplicity, we shall use IVW to represent the random-effects IVW method hereafter.

In Figure 3, we present the percent bias, empirical coverage, and average lengths of 95% confidence intervals of those MR methods in simulated data with a sample size of 5,000 for both the exposure and the outcome. Additional simulation results under a range of sample sizes (n=5,000, 10,000, 20,000, 40,000, 80,000) can be found in Supplementary Figure S1 and Tables S1-S3. When the plurality rule condition holds and no locally invalid IVs exist, MR-SPI has small bias and short confidence interval, and the empirical coverage can attain the nominal level. When locally invalid IVs exist, the standard confidence interval might suffer from finite-sample IV selection error, and thus the empirical coverage is lower than 95% if the sample sizes are not large (e.g., 5,000). In practice, we can perform sensitivity analysis of the causal effect estimate by changing the threshold in the voting step (see STAR Methods and Supplementary Figure S14). If the causal effect estimate is sensitive to the choice of the threshold, then there might exists finite-sample IV selection error. In such cases, the proposed robust confidence interval of MR-SPI can still attain the 95% coverage level and thus is recommended for use. We also examine the performance of MR-SPI in overlapped samples mimicking real data settings with simulation set-up and results given in Supplementary Section S8, and we find that our MR-SPI can still provide valid statistical inference.

**Main Figure 3.**
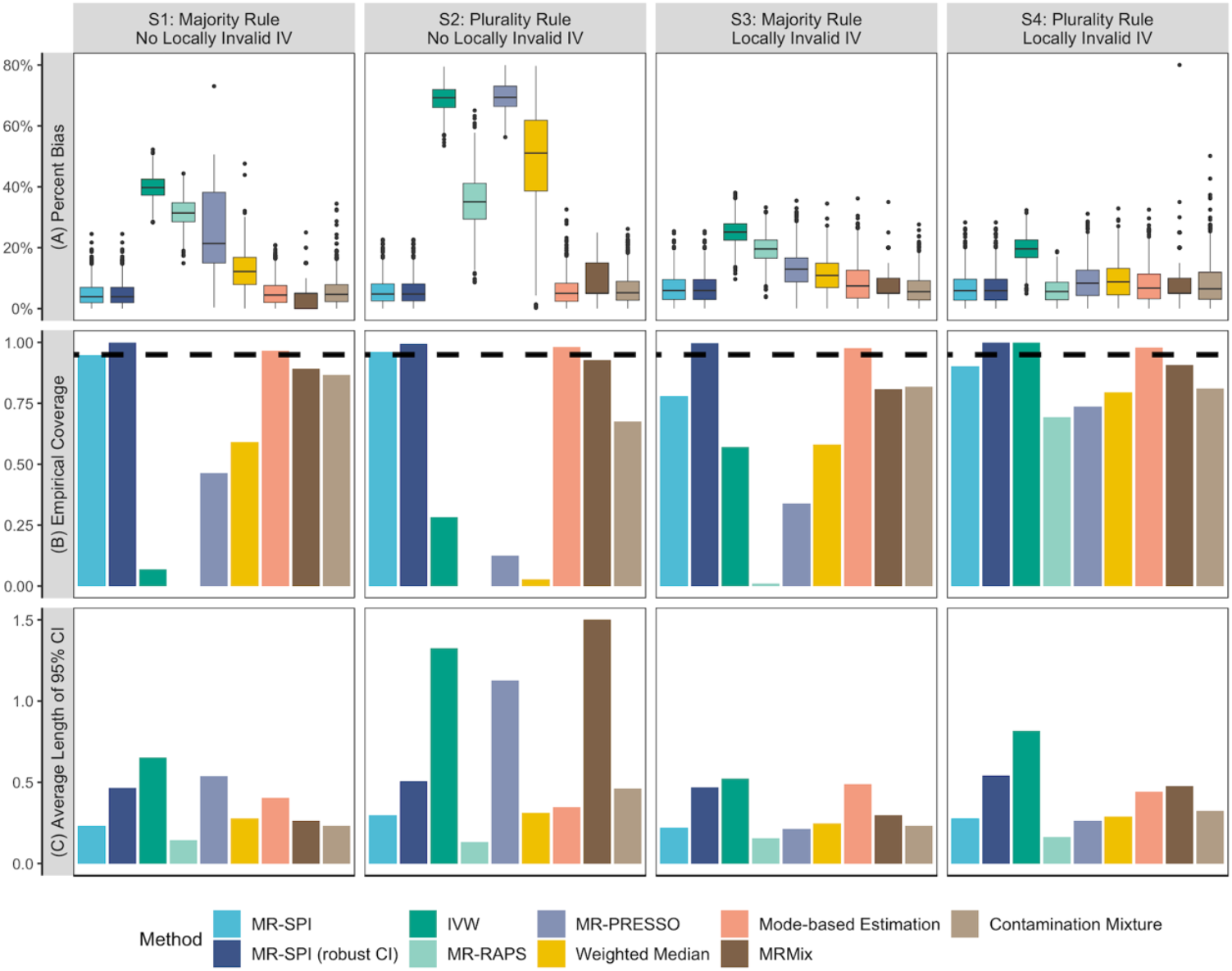
Empirical performance of MR-SPI and the other competing MR methods in simulated data with sample size 5,000. (A) Boxplot of the percent bias in causal effect estimates. (B) Empirical coverage of 95% confidence intervals. The black dashed line in (B) represents the nominal level (95%). (C) Average lengths of 95% confidence intervals.

### 2.4 Identifying plasma proteins associated with the risk of Alzheimer’s disease

Omics MR (xMR) aims to identify omics biomarkers (e.g., proteins) causally associated with complex traits and diseases. In particular, xMR with proteomics data enables the identification of disease-associated proteins, facilitating crucial advancements in disease diagnosis, monitoring, and novel drug target discovery. In this section, we apply MR-SPI to identify putative causal plasma protein biomarkers associated with the risk of Alzheimer’s disease (AD). The proteomics data used in our analysis comprises 54,306 participants from the UK Biobank Pharma Proteomics Project (UKB-PPP)^20^. In the UKB-PPP consortium, up to 22.6 million imputed autosomal variants across 1,463 proteins post quality control were analyzed, discovering 10,248 primary associations through LD (Linkage Disequilibrium) clumping ±1Mb around the significant variants, including 1,163 in the *cis* region and 9,085 in the *trans* region^20^. As described in Sun, et al. ^20^, the following filtering steps are used to retain pQTLs in the UKB-PPP summary level proteomics data: (1)genome-wide significant (*p* -value < 3.40 × 10^−11^), after Bonferroni correction; and (2) independent pQTLs using LD clumping (*r*^2^ < 0.01). Thus, all these candidates pQTL IVs are independent and strongly associated with the proteins. Summary statistics for AD are obtained from a meta-analysis of GWAS studies for clinically diagnosed AD and AD-by-proxy, comprising 455,258 samples in total^50^. For MR method comparison, we analyze 912 plasma proteins that share four or more candidate pQTLs within the summary statistics for AD, because the implementation of MR-PRESSO requires a minimum of four candidate IVs ^29^.

As presented in Figure 4(A), MR-SPI identifies 7 proteins that are significantly associated with AD after Bonferroni correction, including CD33, CD55, EPHA1, PILRA, PILRB, RET, and TREM2. The detailed information of the selected pQTL IVs for these 7 proteins can be found in Supplementary Table S6. Among them, four proteins (CD33, PILRA, PILRB, and RET) are positively associated with the risk of AD while the other three proteins (CD55, EPHA1, and TREM2) are negatively associated with the risk of AD. We also note that some competing MR methods may detect additional proteins, which are likely spurious due to invalid pQTL IVs, as demonstrated in Supplementary Section 11. Previous studies have revealed that some of those 7 proteins and the corresponding protein-coding genes might contribute to the pathogenesis of AD^51-56^, as shown in Supplementary Table S8. For example, it has been found that CD33 plays a key role in modulating microglial pathology in AD, with TREM2 acting downstream in this regulatory pathway^53^. Besides, a recent study has shown that a higher level of soluble TREM2 is associated with protection against the progression of AD pathology^57^. Additionally, RET at mitochondrial complex I is activated during ageing, which might contribute to an increased risk of ageing-related diseases including AD^55^. Using the UniProt database^58^, we also find that genes encoding these 7 proteins are overexpressed in tissues including hemopoietic tissues and brain, as well as cell types including microglial, macrophages and dendritic cells. These findings highlight the potential therapeutic opportunities that target these proteins for the treatment of AD. Furthermore, in the Therapeutic Target Database (TTD)^59^ and DrugBank database^60^, we find existing US Food and Drug Administration (FDA)-approved drugs that target these proteins identified by MR-SPI. For example, gemtuzumab ozogamicin is a drug that targets CD33 and has been approved by FDA for acute myeloid leukemia therapy^61,62^. Besides, pralsetinib and selpercatinib are two RET inhibitors that have been FDA-approved for the treatment of non-small-cell lung cancers^63,64^. Therefore, these drugs might be potential drug repurposing candidates for the treatment of AD.

**Main Figure 4.**
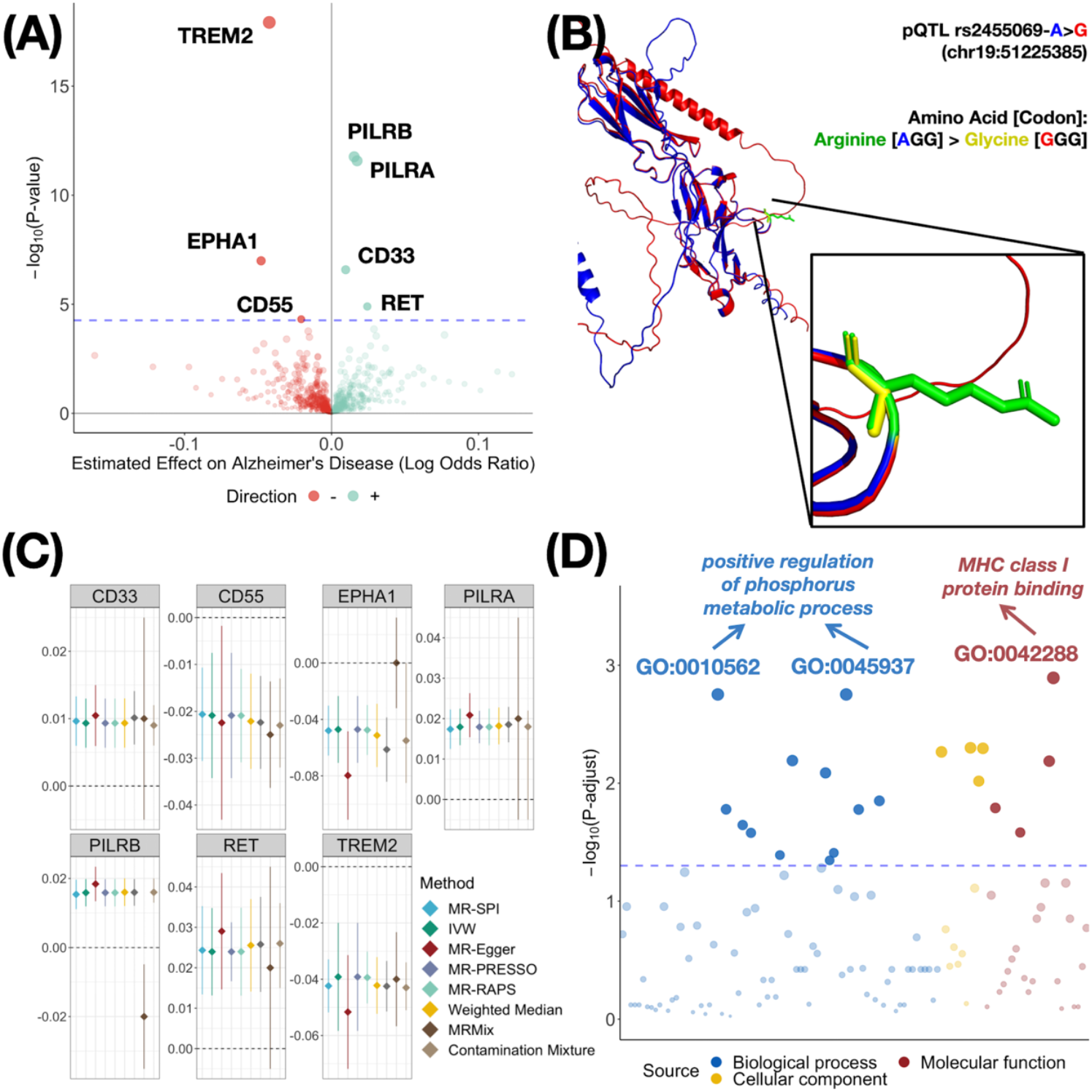
Causal effect estimates, gene ontology analysis and structural alteration prediction for putatively causal proteins. (A) Volcano plot of associations of plasma proteins with Alzheimer’s disease using MR-SPI. The horizontal axis represents the estimated effect size (on the log odds ratio scale), and the vertical axis represents the – log_10_(*p* -value). Positive and negative associations are represented by green and red points, respectively. The size of a point is proportional to − log_10_(*p* -value). The blue dashed line represents the significance threshold using Bonferroni correction (*p* -value < 5.48 × 10^−5^). (B) 3D Structural alterations of CD33 predicted by AlphaFold3 due to missense genetic variation of pQTL rs2455069. The ribbon representation of 3D structures of CD33 with Arginine and Glycine at position 69 are colored in blue and red, respectively. The amino acids at position 69 are displayed in stick representation, with Arginine and Glycine colored in green and yellow, respectively. The predicted template modeling (pTM) yields a score of 0.6 for both structures, which suggests that AlphaFold3 provides good predictions for these two 3D structures. (C) Forest plot of significant associations of proteins with Alzheimer’s disease identified by MR-SPI. Confidence intervals are clipped to vertical axis limits. (D) Bubble plot of GO analysis results using the 7 significant proteins detected by MR-SPI. The horizontal axis represents the *z*-score of the enriched GO term, and the vertical axis represents the − log_10_ (*p*-value) after Bonferroni correction. Each point represents one enriched GO term. The blue dashed line represents the significance threshold (adjusted *p*-value< 0.05 after Bonferroni correction).

In Figure 4(B), we present the 3D structural alterations of CD33 due to missense genetic variation of pQTL rs2455069, as predicted by AlphaFold3^42,43,45^. The 3D structures are shown in blue when the allele is A, and in red when the allele is G at pQTL rs2455069 A/G, which is a cis-SNP located on chromosome 19 (19q13.41) and is selected as a valid IV by MR-SPI. The presence of the G allele at pQTL rs2455069 results in the substitution of the 69th amino acid of CD33, changing it from Arginine (colored in green if the allele is A) to Glycine (colored in yellow if the allele is G), consequently causing a local change in the structure of CD33 (R69G). Previous studies have found that CD33 is overexpressed in microglial cells in the brain^65^, and the substitution of Arginine to Glycine in the 69th amino acid of CD33 might lead to the accumulation of amyloid plaques in the brain^66^, thus the presence of the G allele at pQTL rs2455069 might contribute to an increased risk of AD. We also apply AlphaFold3 to predict the 3D structures of the other proteins that are detected to be significantly associated with AD by MR-SPI, which are presented in Supplementary Figure S16.

In Figure 4(C), we present the point estimates and 95% confidence intervals of the causal effects (on the log odds ratio scale) of these 7 proteins on AD using the other competing MR methods. In Figure 4(C), these proteins are identified by most of the competing MR methods, confirming the robustness of our findings. To the best of our knowledge, there may be two reasons for the differences in results between MRMix and other MR methods for some proteins: (1) MRMix assumes that the pQTL-protein and pQTL-outcome associations follow a bivariate normal-mixture model with four mixture components while the contamination mixture models assume that the ratio estimator follows a normal distribution with two mixture components, and therefore it may be more challenging to obtain reliable causal effect estimates using the MRMix model with a small number of pQTLs per protein; and (2) the default grid search values implemented in the MRMix R package might not be optimal for some proteins. Notably, MR-SPI detects one possibly invalid IV pQTL rs10919543 for TREM2-AD relationship, which is associated with red blood cell count according to PhenoScanner^67^. Red blood cell count is a known risk factor for AD^68,69^, and thus pQTL rs10919543 might exhibit pleiotropy in the relationship of TREM2 on AD. After excluding this potentially invalid IV, MR-SPI suggests that TREM2 is negatively associated with the risk of AD (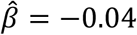, *p*-value= 1.20 × 10^−18^). Additionally, we perform the gene ontology (GO) enrichment analysis using the g:Profiler web server^70^ (https://biit.cs.ut.ee/gprofiler/gost) to gain more biological insights for the 7 proteins identified by MR-SPI, and the results are presented in Figure 4(D) and Supplementary Table S7. After Bonferroni correction, the GO analysis indicates that these 7 proteins are significantly enriched in 20 GO terms, notably, the positive regulation of phosphorus metabolic process and major histocompatibility complex (MHC) class I protein binding. It has been found that increased phosphorus metabolites (e.g., phosphocreatine) are associated with aging, and that defects in metabolic processes for phospholipid membrane function is involved in the pathological progression of Alzheimer’s disease^71,72^. In addition, MHC class I proteins may play a crucial role in preserving brain integrity during post-developmental stages, and modulation of the stability of MHC class I proteins emerges as a potential therapeutic target for restoring synaptic function in AD^73-75^.

## 3. Discussion

In this paper, we develop a novel integrated pipeline that combines our proposed MR-SPI method with AlphaFold3 to identify putative causal protein biomarkers for complex traits/diseases and to predict the 3D structural alterations induced by missense pQTL IVs. Specifically, MR-SPI is an automatic algorithm to select valid pQTL IVs under the plurality rule condition for a specific protein-outcome pair from two-sample GWAS summary statistics. MR-SPI first selects relevant pQTL IVs with strong pQTL-protein associations to minimize weak IV bias, and then applies the proposed voting procedure to select valid pQTL IVs whose ratio estimates are similar to each other. In the possible presence of locally invalid IVs in finite-sample settings, MR-SPI further provides a robust confidence interval constructed by the searching and sampling method^49^, which is immune to finite-sample IV selection error. The valid pQTL IVs selected by MR-SPI serve dual purposes: (1) facilitating more reliable scientific discoveries in identifying putative causal proteins associated with diseases; and (2) shedding new light on the molecular-level mechanism of causal proteins in disease etiology through the 3D structural alterations of mutated proteins induced by missense pQTL IVs. We employ MR-SPI to conduct xMR analysis with 912 plasma proteins using the proteomics data in 54,306 UK Biobank participants and identify 7 proteins significantly associated with the risk of Alzheimer’s disease. The 3D structural changes in these proteins, as predicted by AlphaFold3 in response to missense genetic variations of selected pQTL IVs, offering new insights into their biological functions in the etiology of Alzheimer’s disease. We also found existing FDA-approved drugs that target some of our identified proteins, which provide opportunities for potential existing drug repurposing for the treatment of Alzheimer’s disease. These findings highlight the great potential of our proposed pipeline for identifying protein biomarkers as new therapeutic targets and drug repurposing for disease prevention and treatment.

We emphasize three main advantages of MR-SPI. First, MR-SPI incorporates both proteomics and outcome data to automatically select a set of valid pQTL instruments in genome-wide studies, and the selection procedure does not rely on any additional distributional assumptions on the genetic effects nor require a large number of candidate IVs. Therefore, MR-SPI is the first method to offer such a practically robust approach to selecting valid pQTL IVs for a specific exposure-outcome pair from GWAS studies for more reliable MR analyses, which is especially advantageous in the presence of wide-spread horizontal pleiotropy and when only a small number of candidate IVs are available in xMR studies. While our real data application specifically focuses on the identification of putative causal protein biomarkers for Alzheimer’s disease through the integration of MR-SPI with AlphaFold3, it’s important to highlight that MR-SPI holds broader applicability in elucidating causal relationships across complex traits and diseases. For additional data analysis results and insights into the utility of MR-SPI in this context, please refer to Supplementary Sections S9 and S10. Second, we propose a robust confidence interval for the causal effect using the searching and sampling method, which is immune to finite-sample IV selection error. Therefore, when locally invalid IVs are incorrectly selected, MR-SPI can still provide reliable statistical inference for the causal effect using the proposed robust confidence interval. Third, MR-SPI is computationally efficient. The average computation time for constructing the standard CI and the robust CI with 20 candidate IVs is 0.02 seconds and 10.60 seconds, respectively, using a server equipped with an Intel Xeon Silver 4116 CPU and 64 GB RAM memory.

In conclusion, MR-SPI is a powerful tool for identifying putative causal protein biomarkers for complex traits and diseases. The integration of MR-SPI with AlphaFold3 as a computationally efficient pipeline can further predict the 3D structural alterations caused by missense pQTL IVs, improving our understanding of molecular-level disease mechanisms. Therefore, our pipeline holds promising implications for drug target discovery, drug repurposing, and therapeutic development.

### Limitations of the Study

MR-SPI has some limitations. First, MR-SPI uses independent pQTLs as candidate IVs after LD clumping, which might exclude strong and valid pQTL IVs. We plan to extend MR-SPI to include correlated pQTLs with arbitrary LD structure to increase statistical power. Second, the proposed robust confidence interval is slightly more conservative, which is the price to pay for the gained robustness to finite-sample IV selection error. We plan to construct less conservative confidence intervals with improved power to detect more putative causal proteins. Third, we will incorporate colocalization analysis^76-79^ into our pipeline to better understand the shared genetic architecture between proteins and disease outcomes when unfiltered GWAS summary statistics are available in future studies.

## Data Availability

All data produced in the present study are available upon reasonable request to the authors.

## 4. Resource Availability

### Lead Contact

Further information and requests for resources and reagents should be directed to and will be fulfilled by the lead contact, Zhonghua Liu

## Materials Availability

The materials that support the findings of this study are available from the corresponding authors upon reasonable request. Please contact the lead contact, Zhonghua Liu for additional information.

## Data and Code Availability

All the GWAS data analyzed are publicly available with the following URLs:

- GWAS for Alzheimer’s disease: https://ctg.cncr.nl/software/summary_statistics;
- UK Biobank proteomics data: https://www.biorxiv.org/content/10.1101/2022.06.17.496443v1.supplementary-material

The R package **MR.SPI** is publicly available at https://github.com/MinhaoYaooo/MR-SPI.

## 5. Acknowledgments

We thank the UK Biobank Pharma Proteomics Project (http://ukb-ppp.gwas.eu/) for providing the proteomics data. We thank the Center for Neurogenomics and Cognitive Research (CNCR, https://cncr.nl/) for providing GWAS summary statistics for Alzheimer’s disease.

## 6. Author Contributions

Conceptualization, Zijian Guo and Zhonghua Liu; methodology, Minhao Yao, Zijian Guo and Zhonghua Liu; software, Minhao Yao; formal analysis: Minhao Yao; writing (original draft), Minhao Yao, Zijian Guo and Zhonghua Liu; writing (review & editing), Minhao Yao, Gary W. Miller, Badri N. Vardarajan, Andrea A. Baccarelli, Zijian Guo and Zhonghua Liu; supervision, Zijian Guo and Zhonghua Liu.

## STAR Methods

### Key resources table

STAR Methods Key resources table
| REAGENT or RESOURCE | SOURCE | IDENTIFIER |
| --- | --- | --- |
| **Deposited data** |  |  |
| UK Biobank proteomics data | Sun, et al.<sup>20</sup> | http://ukb-ppp.gwas.eu/ |
| Genome-wide association studies of Alzheimer's disease | Jansen, et al.<sup>50</sup> | https://www.nature.com/articles/s41588-018-0311-9 |
| **Software and algorithms** |  |  |
| AlphaFold3 | Abramson, et al.<sup>45</sup> | https://alphafoldserver.com |
| g:Profiler | Raudvere, et al.<sup>70</sup> | https://biit.cs.ut.ee/gprofiler/gost |
| MendelianRandomization | Yavorska and Burgess<sup>80</sup> | https://github.com/cran/MendelianRandomization |
| MRMix | Qi and Chatterjee<sup>32</sup> | https://github.com/gqi/MRMix |
| MR-PRESSO | Verbanck, et al.<sup>29</sup> | https://github.com/rondolab/MR-PRESSO |
| MR-RAPS | Zhao, et al.<sup>16</sup> | https://github.com/qingyuanzhao/mr.raps |
| MR-SPI | This study | https://github.com/MinhaoYaooo/MR-SPI |
| PLINK | Purcell, et al.<sup>81</sup> | https://www.cog-genomics.org/plink/1.9 |
| PyMol | PyMOL Molecular Graphics | https://pymol.org |
|  | System |  |
| R | The R Foundation for Statistical Computing | https://www.r-project.org/ |

### Method details

#### Two-sample GWAS summary statistics

Suppose that we obtain *p* independent pQTLs ***Z*** = (*Z*^1^, ⋯, *Z*_*p*_)^⊤^ by using LD clumping that retains one representative pQTL per LD region^81^. We also assume that the pQTLs are standardized^82^ such that 𝔼 *Z*_*j*_ = 0 and var (*Z*_*j*_)= 1 for 1 ≤ *j* ≤ *p*. Let *D* denote the exposure and *Y* denote the outcome. We assume that *D* and *Y* follow the exposure model *D* = ***Z***^⊤^***γ*** + *δ* and the outcome model *Y* = *Dβ* + ***Z***^⊤^ ***π*** + *e*, respectively, where *β* represents the causal effect of interest, ***γ*** = (*γ*, ⋯, *γ*)^⊤^ represents the IV strength, and ***π*** = (*π*_1_⋯,*π*_*p*_)^⊤^ @encodes the violation of assumptions (A2) and (A3)^83,84^. If assumptions (A2) and (A3) hold for pQTL *j*, then *π*_*j*_ = 0 and otherwise *π*_*j*_ ≠ 0 (see Supplementary Section S1 for details). The error terms *δ* and *e* with respective variances 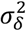 and 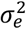 are possibly correlated due to unmeasured confounding factors. By plugging the exposure model into the outcome model, we obtain the reduced-form outcome model *Y* = ***Z***^⊤^ (*β****γ*** + ***π***) + *∈*, where *∈* = *βδ* + *e*. Let **Γ** = (Γ_1_, ⋯, Γ_*p*_)^⊤^ denote the pQTL-outcome associations, then we have **Γ** = *β****γ*** + ***π***. If *γ*_*j*_ ≠ 0, then pQTL *j* is called a relevant IV. If both *γ*_*j*_ ≠ 0 and *π*_*j*_ = 0, then pQTL *j* is called a valid IV. Let 𝒮 = {*j*: *γ*_*j*_ ≠ 0,1 ≤ *j* ≤ *p*} denote the set of all relevant IVs, and 𝒱 = {*j*: *γ*_*j*_ ≠ 0 and *π*_*j*_ = 0,1 ≤ *j* ≤ *p*} denote the set of all valid IVs. The majority rule condition can be expressed as 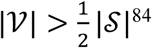, and the plurality rule condition can be expressed as |𝒱| > max_*c*≠*o*_|{*j* ∈ 𝒮:*π*_*j*_/*γ*_*j*_ = *c*}|^83^. If the plurality rule condition holds, then valid IVs with the same ratio of pQTL-outcome effect to pQTL-protein effect will form a plurality. Based on this key observation, our proposed MR-SPI selects the largest group of pQTLs as valid IVs with similar ratio estimates of the causal effect using a voting procedure described in detail in the next subsection.

Let 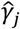 and 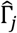 be the estimated marginal effects of pQTL *j* on the protein and the outcome, and 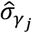 and 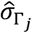 be the corresponding estimated standard errors respectively. Let 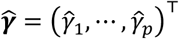 and 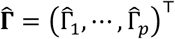 denote the vector of estimated pQTL-protein and pQTL-outcome associations, respectively. In the two-sample setting, the summary statistics 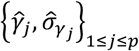and 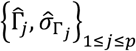 are calculated from two non-overlapping samples with sample sizes *n*_1_ and *n*_2_ respectively. When all the pQTLs are independent of each other, the joint asymptotic distribution of 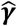 and 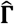 is

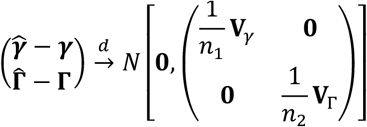

where the diagonal entries of **V**_*γ*_ and **V**_Γ_ are 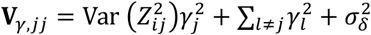 and 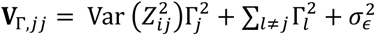, respectively, and the off-diagonal entries of **V**_*γ*_ and **V**_Γ_ are 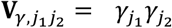 and 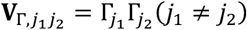, respectively. The derivation of the limit distribution can be found in Supplementary Section S2. Therefore, with the summary statistics of the protein and the outcome, we estimate the covariance matrices 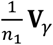 and 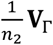 as:

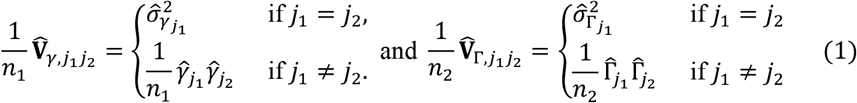

After obtaining 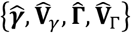, we then perform the proposed IV selection procedure as illustrated in Figure 2 in the main text.

#### Selecting valid instruments by voting

The first step of MR-SPI is to select relevant pQTLs with large IV strength using proteomics data. Specifically, we estimate the set of relevant IVs 𝒮 by:

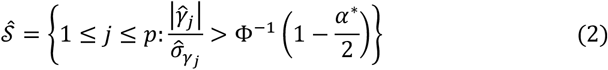

where 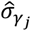 is the standard error of 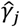 in the summary statistics, Φ^-1^(⋅) is the quantile function of the standard normal distribution, and *α*^*^ is the user-specified threshold with the default value of 1 × 10^−6^. This step is equivalent to filtering the pQTLs in the proteomics data with *p*-value < *α*^*^, and is adopted by most of the current two-sample MR methods to select (relevant) genetic instruments for downstream MR analysis. Note that the selected pQTL instruments may not satisfy the IV independence and exclusion restriction assumptions and thus maybe invalid. In contrast, our proposed MR-SPI further incorporates the outcome data to automatically select a set of valid genetic instruments from 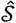 for a specific protein-outcome pair.

Under the plurality rule condition, valid pQTL instruments with the same ratio of pQTL-outcome effect to pQTL-protein effect (i.e., Γ_*j*_/*γ*_*j*_) will form a plurality and yield “similar” ratio estimates of the causal effect. Based on this key observation, MR-SPI selects a plurality of relevant IVs whose ratio estimates are “similar” to each other as valid IVs. Specifically, we propose the following two criteria to measure the similarity between the ratio estimates of two pQTLs *j* and *k* :

**C1**: We say the *k*th pQTL “votes for” the *j*th pQTL to be a valid IV if, by assuming the *j*th pQTL is valid, the *k*th pQTL’s degree of violation of assumptions (A2) and (A3) is smaller than a threshold as in equation (4);

**C2**: We say the ratio estimates of two pQTLs *j* and *k* are “similar” if they mutually vote for each other to be valid IVs.

The ratio estimate of the *j*th pQTL is defined as 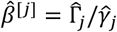. By assuming the *j*th pQTL is valid, the plug-in estimate of the *k*th pQTL’s degree of violation of (A2) and (A3) can be obtained by

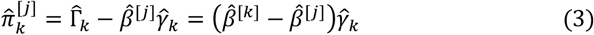

as we have Γ_*k*_ = *βγ*_*k*_ + *π*_*k*_ for the true causal effect *β*, and 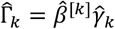 for the ratio estimate 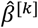 of the *k* th pQTL. From equation (3), 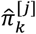 has two noteworthy implications. First, 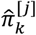 measures the difference between the ratio estimates of pQTLs *j* and *k* (multiplied by the *k*th pQTL-protein effect estimate 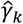), and a small 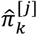 implies that the difference scaled by 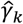 is small. Second, 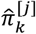 represents the *k*th IV’s degree of violation of assumptions (A2) and (A3) by regarding the *j*th pQTL’s ratio estimate 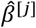 as the true causal effect, thus a small 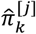 implies a strong evidence that the *k*th IV supports the *j*th IV to be valid. Therefore, we say the *k*th IV votes for the *j*th IV to be valid if:

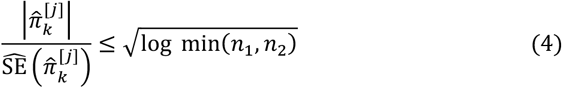

where 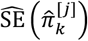 is the standard error of 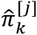, which is given by:

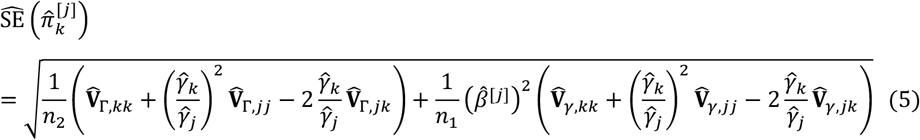

and the term 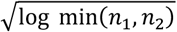 in equation (4) ensures that the violation of (A2) and (A3) can be correctly detected with probability one as the sample sizes go to infinity, as shown in Supplementary Section S3.

For each relevant IV in 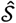, we collect all relevant IVs’ votes on whether it is a valid IV according to equation (4). Then we construct a voting matrix 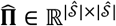 to summarize the voting results and evaluate the similarity of two pQTLs’ ratio estimates according to criterion C2. Specifically, we define the (*k, j*) entry of 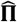 as:

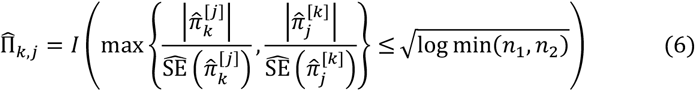

where *I*(⋅) is the indicator function such that *I*(*A*) = 1 if event *A* happens and *I*(*A*) = 0 otherwise. From equation (6), we can see that the voting matrix 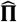 is symmetric, and the entries of 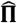 are binary: 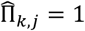 represents pQTLs *j* and *k* vote for each other to be a valid IV, i.e., the ratio estimates of these two pQTLs are close to each other; 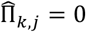 represents that they do not. For example, in Figure 2, 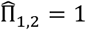 since the ratio estimates of pQTLs 1 and 2 are similar, while 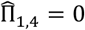 because the ratio estimates of pQTLs 1 and 4 differ substantially, as pQTLs 1 and 4 mutually “vote against” each other to be valid according to equation (4).

After constructing the voting matrix 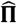, we select the valid IVs by applying majority/plurality voting or finding the maximum clique of the voting matrix^47^. Let 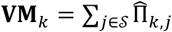 be the total number of pQTLs whose ratio estimates are similar to pQTL *k*. For example, **VM**_1_ = 3 in Figure 2, since three pQTLs (including pQTL 1 itself) yield similar ratio estimates to pQTL 1 according to criterion **C2**. A large **VM**_*k*_ implies strong evidence that pQTL *k* is a valid IV, since we assume that valid IVs form a plurality of the relevant IVs. Let 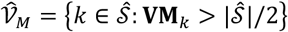 denote the set of IVs with majority voting, and 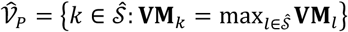 denote the set of IVs with plurality voting, then the union 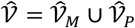 can be a robust estimate of 𝒱 in practice. Alternatively, we can also find the maximum clique in the voting matrix as an estimate of 𝒱. A clique in the voting matrix is a group of IVs who mutually vote for each other to be valid, and the maximum clique is the clique with the largest possible number of IVs^47^.

#### Estimation and inference of the causal effect

After selecting the set of valid pQTL instruments 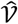, the causal effect *β* is estimated by

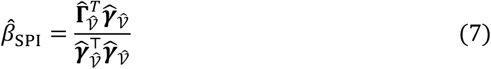

where 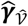 and 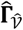 are the estimates of pQTL-protein associations and pQTL-outcome associations of the selected valid IVs in 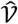, respectively. The MR-SPI estimator in equation (7) is the regression coefficient obtained by fitting a zero-intercept ordinary least squares regression of 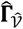 on 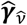. Since the pQTLs are standardized, the genetic associations 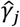 and 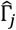 are scaled by 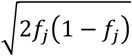 (compared to the genetic associations calculated using the unstandardized pQTLs, denoted by 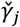 and 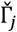), where *f*_*j*_ is the minor allele frequency of pQTL *j*. As *f*_*j*_(1 − *f*_*j*_) is approximately proportional to the inverse variance of 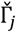 when each pQTL IV explains only a small proportion of variance in the outcome^85^, the MR-SPI estimator of the causal effect in equation (7) is approximately equal to the inverse-variance weighted estimator^27^ calculated with 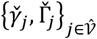.

Let *α* ∈ (0,1) be the significance level and *z*_1-*α*/2_ be the (1 − *α*/2)-quantile of the standard normal distribution, then the (1 − *α*) confidence interval for *β* is given by:

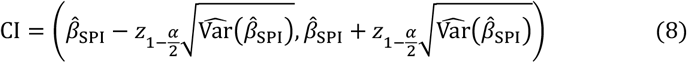

where 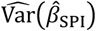 is the estimated variance of 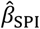, which can be found in Supplementary Section S4. As 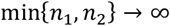, we have 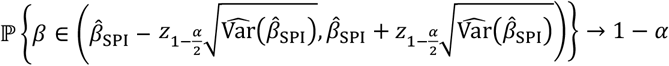 under the plurality rule condition, as shown in Supplementary Section S5. Hence, MR-SPI provides a theoretical guarantee for the asymptotic coverage probability of the confidence interval under the plurality rule condition. We summarize the proposed procedure of selecting valid IVs and constructing the corresponding confidence interval by MR-SPI in Algorithm 1.

#### A robust confidence interval via searching and sampling

In finite-sample settings, the selected set of relevant IVs 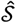 might include some invalid IVs whose degrees of violation of (A2) and (A3) are small but nonzero, and we refer to them as “locally invalid IVs”^49^. When locally invalid IVs exist and are incorrectly selected into 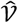, the confidence interval in equation (8) becomes unreliable, since its validity (i.e., the coverage probability attains the nominal level) requires that the invalid IVs are correctly filtered out. In practice, we can multiply the threshold 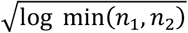 in the right-hand side of equation (4) by a scaling factor *η* to examine whether the confidence interval calculated by equation (8) is sensitive to the choice of the threshold. If the confidence interval varies substantially to the choice of the scaling factor *η*, then there might exist finite-sample IV selection error especially with locally invalid IVs. We demonstrate this issue with two numerical examples presented in Supplementary Figure S14. Supplementary Figure S14(a) shows an example in which MR-SPI provides robust inference across different values of the scaling factor, while Supplementary Figure S14(b) shows an example that MR-SPI might suffer from finite-sample IV selection error, as the causal effect estimate and the corresponding confidence interval are sensitive to the choice of the scaling factor *η*. This issue motivates us to develop a more robust confidence interval.

To construct a confidence interval that is robust to finite-sample IV selection error, we borrow the idea of searching and sampling^49^, with main steps described in Supplementary Figure S17. The key idea is to sample the estimators of ***γ*** and **Γ** repeatedly from the following distribution:

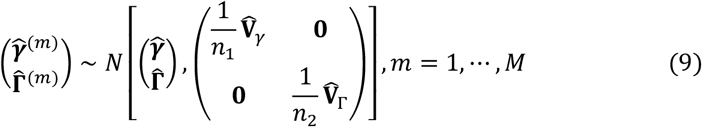

where *M* is the number of sampling times (by default, we set *M* = 1,000). Since 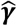 and 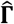 follow distributions centered at ***γ*** and **Γ**, there exists *m*^*^ such that 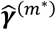 and 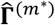 are close enough to the true values ***γ*** and **Γ** when the number of sampling times *M* is sufficiently large, and thus the confidence interval obtained by using 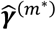 and 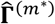 instead of 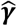 and 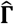 might have a larger probability of covering *β*.

For each sampling, we construct the confidence interval by searching over a grid of *β* values such that more than half of the selected IVs in 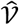 are detected as valid. As for the choice of grid, we start with the smallest interval [*L, U*] that contains all the following intervals:

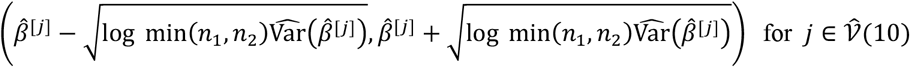

where 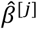 is the ratio estimate of the *j* th pQTL IV, 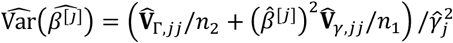 is the variance of 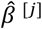, and 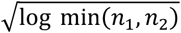 serves the same purpose as in equation (4). Then we discretize [*L, U*] into 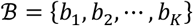 as the grid set such that *b*_1_ = *L, b*_*K*_ = *U* and 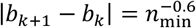 for 1 ≤ *k* ≤ *K* − 2, where *n*_min_ = min(*n*_1_, *n*_2_). We set the grid size 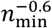 so that the error caused by discretization is smaller than the parametric rate 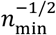.

For each grid value *b* ∈ ℬ and sampling index 1 ≤ *m* ≤ *M*, we propose an estimate of *π*_*j*_ by 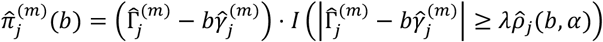 for 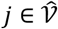,where 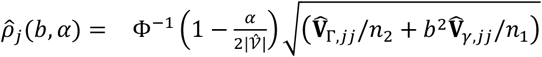 is a data-dependent threshold, Φ^-1^(⋅) is the inverse of the cumulative distribution function of the standard normal distribution, *α* ∈ (0,1) is the significance level, and 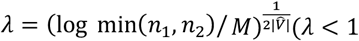 when *M* is sufficiently large) is a scaling factor to make the thresholding more stringent so that the confidence interval in each sampling is shorter, as we will show shortly. Here, 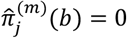 indicates that the *j*th pQTL is detected as a valid IV in the *m* th sampling if we take 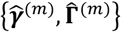 as the estimates of genetic associations and *b* as the true causal effect. Let 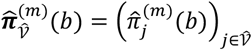, then we construct the *m* th sampling’s pseudo confidence interval pCI^(*m*)^ by searching for the smallest and largest *b* ∈ ℬ such that more than half of pQTLs in 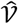 are detected to be valid. Define 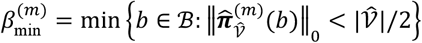 and 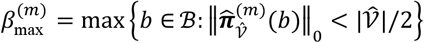, then the *m* th sampling’s pseudo confidence interval is constructed as 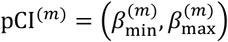.

From the definitions of 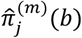 and 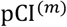, we can see that, when *λ* is smaller, there will be fewer pQTLs in 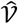 being detected as valid for a given *b* ∈ ℬ, which leads to fewer *b* ∈ ℬ satisfying 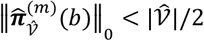, thus the pseudo confidence interval in each sampling will be shorter. If there does not exist *b* ∈ ℬ such that the majority of IVs in ℬ are detected as valid, we set pCI^(*m*)^ = ∅. Let ℳ = {1 ≤ *m* ≤ *M*: pCI^(*m*)^ ≠ ∅} denote the set of all sampling indexes corresponding to non-empty searching confidence intervals, then the proposed robust confidence interval is given by:

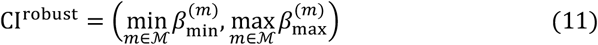

We summarize the procedure of constructing the proposed robust confidence interval in Algorithm 2.

#### Simulation settings

We set the number of candidate IVs *p* = 10, as the average number of candidate pQTL IVs for the plasma proteins in the UK Biobank proteomics data is around 7.4. We set the sample sizes *n*_1_ = *n*_2_ = 5,000, 10,000, 20,000, 40,000, or 80,000. We generate the *j* th genetic instruments *Z*_*j*_ and *X*_*j*_ independently from a binomial distribution Bin (2, *f*_*j*_), where *f*_*j*_ ∼ *U*(0.05,0.50) is the minor allele frequency of pQTL *j*. Then we generate the protein level 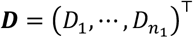 and the outcome 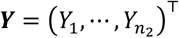 according to the exposure model and the outcome model, respectively. Finally, we calculate the genetic associations and their corresponding standard errors for the protein and the outcome, respectively. As for the parameters, we fix the causal effect *β* = 1, and we consider 4 settings for *γ* ∈ ℝ^*p*^ and *π* ∈ ℝ^*p*^ :

(**S1**): set ***γ*** = 0.2 ⋅ (**1**_5_, −**1**_5_)^⊤^ and ***π*** = 0.2 ⋅ (**0**_6_, **1**_4_)^⊤^.

(**S2**): set ***γ*** = 0.2 ⋅ (**1**_5_, −**1**_5_)^⊤^ and ***π*** = 0.2 ⋅ (**0**_4_, **1**_3_, −**1**_3_)^⊤^.

(**S3**): set ***γ*** = 0.2 ⋅ (**1**_5_, −**1**_5_)^⊤^ and ***π*** = 0.2 ⋅ (**0**_6_, **1**_2_, 0.25,0.25)^⊤^.

(**S4**): set ***γ*** = 0.2 ⋅ (**1**_5_, −**1**_5_)^⊤^ and ***π*** = 0.2 ⋅ (**0**_4_, **1**_2_, 0.25, **1**_2_, −0.25)^⊤^.

Settings (**S1**) and (**S3**) satisfy the majority rule condition, while (**S2**) and (S4) only satisfy the plurality rule condition. In addition, (**S3**) and (**S4**) simulate the cases where locally invalid IVs exist, as we shrink some of the pQTLs’ violation degrees of assumptions (A2) and (A3) down to 0.25 times in these two settings. In total, we run 1,000 replications in each setting.

#### Implementation of existing MR methods

We compare the performance of MR-SPI with eight other MR methods in simulation studies and real data analyses. These methods are implemented as follows:

- Random-effects IVW, MR-Egger, the weighted median method, the mode-based estimation and the contamination mixture method are implemented in the R package “MendelianRandomization” (https://github.com/cran/MendelianRandomization). The mode-based estimation is run with “iteration = 1000”. All other methods are run with the default parameters.
- MR-PRESSO is implemented in the R package “MR-PRESSO” (https://github.com/rondolab/MR-PRESSO) with outlier test and distortion test.
- MR-RAPS is performed using the R package “mr.raps” (https://github.com/qingyuanzhao/mr.raps) with the default options.
- MRMix is run with the R package “MRMix” (https://github.com/gqi/MRMix) using the default options.

##### Algorithm 1: Selecting pQTL IVs and Performing Causal Inference by MR-SPI

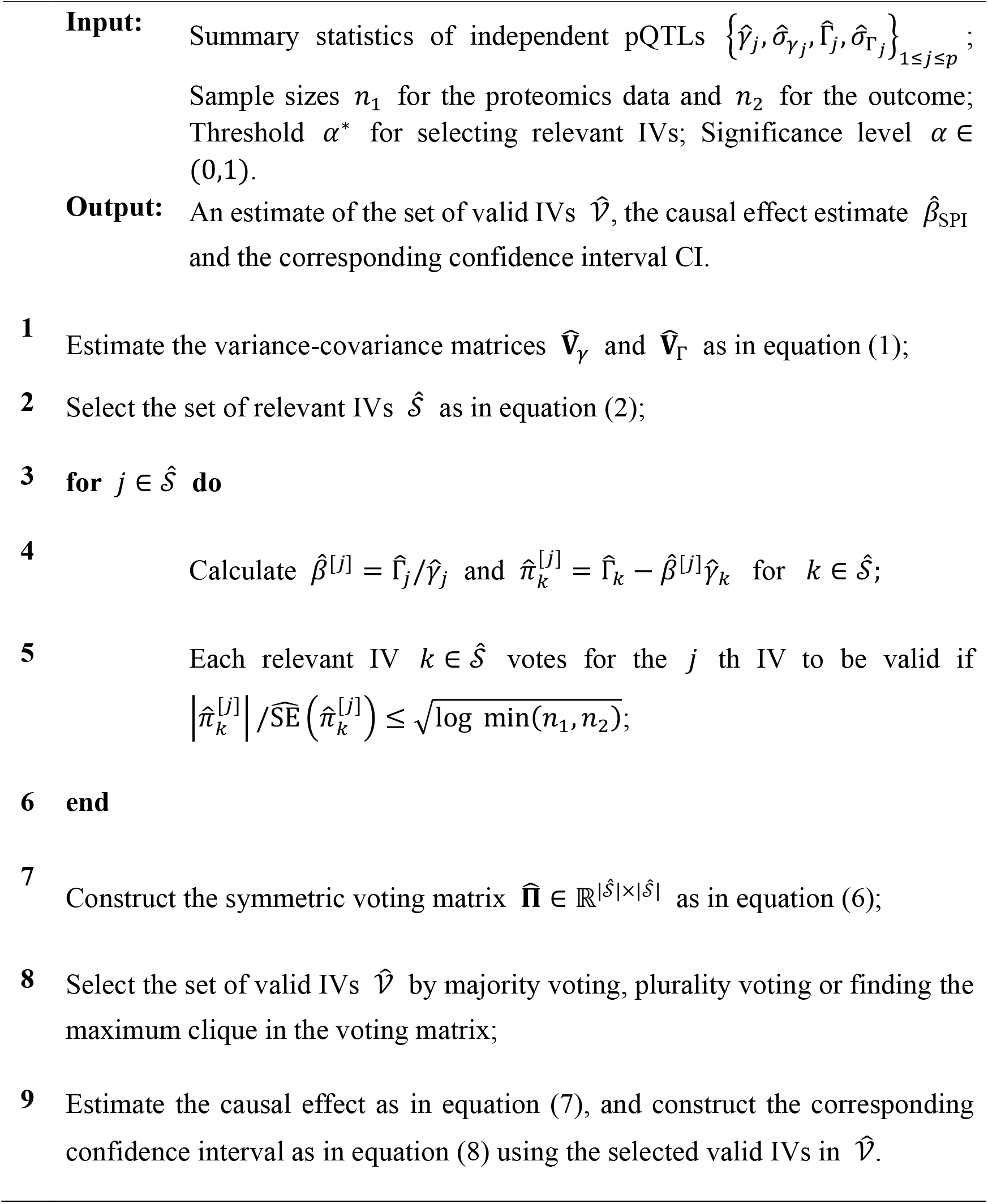

##### Algorithm 2: Constructing A Robust Confidence Interval via Searching and Sampling

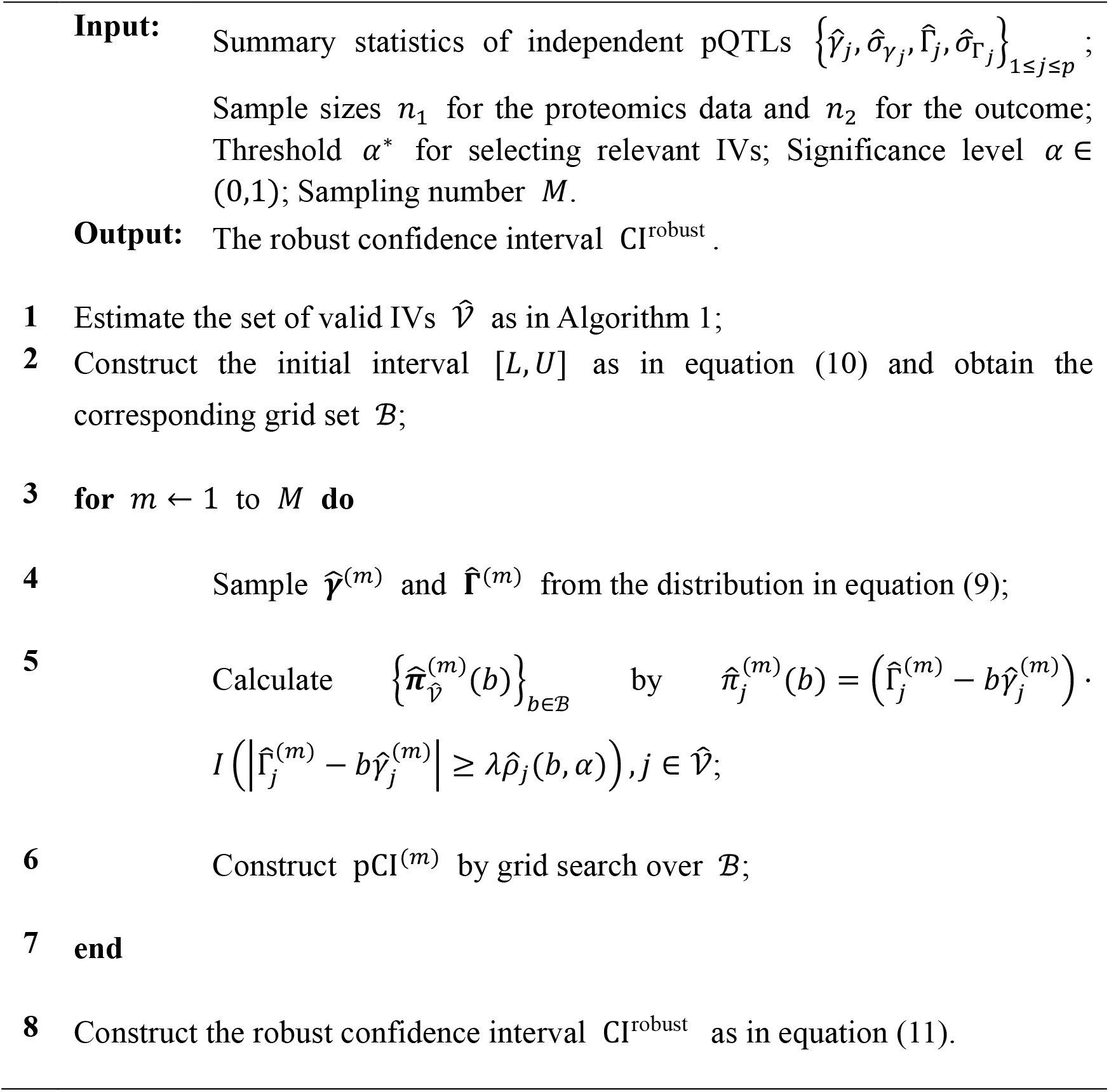

